# Status, determinants and risk factors of all-cause dementia in South Asia: Findings from a preliminary analysis of global health data

**DOI:** 10.1101/2024.03.06.24303854

**Authors:** Dushani L. Palliyaguru, Nipuni Palliyaguru, Camila Vieira Ligo Teixeira, Nicole M. Armstrong, Sanka Liyanage, Upul Senarath, Carukshi Arambepola, Saroj Jayasinghe, Chamila Dalpatadu

## Abstract

South Asia is one of the most populous regions of the world. At present, South Asians make up more than 25% of the world’s population but little effort has been dedicated to studying trends and etiological factors driving aging and aging-related conditions in this part of the world. Even less characterization has been done on brain-related conditions, particularly all-cause dementia in South Asia. To address this, we examined data from the US Census Bureau, World Health Organization Global Dementia Observatory and the Global Burden of Disease (GBD) 2019 Study, and conducted a comparative analysis of population statistics, dementia as a priority health area, and dementia incidence rates, death rates and disability-adjusted life years (DALYs) in 8 South Asian countries – Afghanistan, Bangladesh, Bhutan, India, Maldives, Nepal, Pakistan and Sri Lanka. Our analysis reiterated that there are limited resources dedicated to dementia in this region. Sri Lanka and Afghanistan had the highest dementia crude rates and age-standardized rates, respectively. The burden of dementia in some South Asian countries was comparable to estimated global averages, and was largely driven by population aging. Analyses of available data on known biological, behavioral, environmental and disease risk factors, highlighted the role of metabolic risk factors such as high fasting blood glucose. Here, we also underscore the critical need for future follow-up longitudinal studies focusing on brain aging and dementia and describe a roadmap for designing them - taking into account cultural, economic and public health dynamics that may be uniquely applicable to countries in South Asia.

**KEY MESSAGE:** Very few resources are dedicated to dementia care and research in South Asia, in spite of high burden in some countries. We examine publicly available global health data to highlight that metabolic dysfunction plays a critical role in dementia risk and also detail considerations for future follow-up programs aiming to study dementia in South Asia.

## INTRODUCTION

Aging results in multiple morphological and biochemical changes in the central and peripheral nervous systems. In some individuals, these changes may ultimately lead to dementia [1] - characterized by loss of function in brain regions responsible for visuo-spatial memory, orientation, thinking, learning, language skills and reasoning [2]. While aging is the strongest risk factor for dementia, the likelihood of developing the disease is exacerbated by other behavioral, biological, genetic and environmental factors [3]. Modifiable risk factors of dementia include low physical activity, obesity, tobacco smoking, alcohol consumption and unhealthy eating habits. Dementia is caused by and includes several conditions [4], e.g., Alzheimer’s disease, Vascular disease, Lewy body disease, Frontotemporal dementia, Alcohol-related dementia, HIV-related dementia and Brain injury-related dementia. This debilitating disease is characterized by progressive loss of cognitive capacity which leads to increased dependency, reduced quality of life and mortality [5] [6]. There are no cures for this disease, and currently, most treatments only focus on symptom management [7].

The Global Burden of Disease (GBD) 2019 study estimated staggering increases in dementia rates worldwide over the next few decades [8]. Out of these, low-and-middle-income (LMIC) countries may experience very high financial burden due to dementia [9]. While population growth and aging play a substantial role in this, the poor control of risk factors stemming from lack of awareness, absence of resources and limited research efforts on aging populations may be contributing to this trend [10][11]. Much remains unknown about regional and country-specific dementia statistics and determinants – which are most relevant to policy related decision-making.

South Asia is currently the most highly and densely populated region of the world. It is estimated that over 2 billion people live in South Asia, which accounts for more than 25% of the world’s population at present. Despite significant economic advancements made in some countries, South Asia has one of the lowest Gross Domestic Product (GDP) per capita in the world ($2000) [12]. A significant proportion of these people are part of a rapidly aging population [13] where one in five adults will be over the age of 65 by 2050 in South Asia. Many South Asian countries are still deeply plagued by communicable diseases. However, with rapidly increasing industrialization, many regions of South Asia are quickly becoming epicenters for aging-related chronic diseases, e.g., heart disease, diabetes mellitus and cancer [14]. Previous studies have already identified risk factors for chronic diseases in South Asian populations, e.g., hypertension, hyperglycemia and hypercholesterolemia [15][16][17], which may co-occur in individuals as they age. Other studies have also identified genetic factors that appear to strongly contribute to disease phenotypes observed amongst South Asian populations [18][19]. Yet, so much more remain unknown about biological and demographic factors driving health in this part of the world.

According to early reports, the burden of dementia in South Asia is substantial but health policy, awareness, and research areas do not adequately address this pressing need. Even though South Asian countries are typically aggregated in global health classifications, there is high variability and diversity in individual health status, determinants and trends. Therefore, it is important to examine country-specific dementia data. Such an approach can also lead to the identification of potential hotspots for disease and its burden which can help with targeted disease mitigation efforts. Importantly, it can also help quantify the burden of dementia at different levels, e.g., individuals, caregivers, health sectors and community. In the present study, we examined dementia-related data from multiple public sources to explore and compare the status of the disease as well as risk factors that may be contributing to disease outcomes between different countries.

## METHODS

### Definitions

South Asia was defined according to the World Bank classification of countries which includes Afghanistan, Bangladesh, Bhutan, India, Maldives, Nepal, Pakistan and Sri Lanka [20].

For the present study, the standard definitions of dementia were used as per the GBD 2019 study [21]. It used the Diagnostic and Statistical Manual of Mental Disorders (DSM)-III, DSM-III-R, DSM-IV, or DSM-5, or the International Classification of Diseases (ICD)-8, ICD-9, ICD-10 in representative surveys for dementia cases. This is based on the WHO definition of dementia - an umbrella term used to refer to several diseases affecting memory, other cognitive abilities, and behavior that interfere with a person’s ability to maintain their activities of daily living. This definition includes Alzheimer’s disease, vascular dementia, dementia with Lewy bodies, frontotemporal dementia and dementia that develops because of HIV, alcohol abuse, brain injury and nutritional deficiencies.

### Datasets

#### US Census data

Global demographic data were accessed through the international database of the census.gov website [22]. With this publicly available database, demographic measures of 200 countries can be accessed. Data from censuses, administrative records, vital statistics and National statistical offices of countries are compiled here. For the present analysis, we examined total population, population density, life expectancy at birth, dependency ratio for 65+ and percentage of 65+ year-old people in total population.

#### WHO Global Dementia Observatory data

Through this data and knowledge exchange platform [23], access to key dementia data from 62 participating countries on policies, service delivery, information and research is provided. For the present analysis, we examined whether each South Asian country has a dementia national plan, contributes to the dementia observatory and the availability and status of dementia reporting in the past two years (2017-2019).

#### Global Burden of Disease (GBD) 2019

Dementia incidence, death and disability-adjusted life years (DALY) rates data as well as risk factor data used in the current study were obtained from the Global Health Data Exchange (GHDx) Query Tool [24]. Detailed methods of the GBD 2019 Study have been published previously [21]. Original sources of GBD data include administrative sources, vital registries, disease registries, census/surveys, surveillance systems, hospital records, environmental and geospatial monitors, and scientific literature. Original data from multiple sources are compiled, corrected for known biases, entered into a database and passed through statistical modeling pipelines (e.g., Space-time and Gaussian Process Regression) that allow for comparison between countries. Modeling and estimate methods are used when reliable statistics cannot be obtained or are missing. Several improvements to the GBD study’s data collection and compilation methods have been made since its inception [25]. Since data are publicly available estimates from deidentified sources, informed consent was not applicable. GBD study complies with the Guidelines for Accurate and Transparent Health Estimates Reporting (GATHER) [26].

### Risk factors

Six behavioral, metabolic and environmental risk factors (low physical activity, tobacco use, alcohol use, obesity, hypertension, and air pollution) and three disease risk factors (diabetes mellitus, depression, hearing loss) were examined through the Global Health Data Exchange (GHDx) Query Tool [24]. These were selected based on current knowledge and prior literature on dementia risk factors [27][28][29][30][31][32][33]. Tobacco included smoking, chewing tobacco and secondhand smoking. Obesity and hypertension were examined through high body mass index and high systolic blood pressure, respectively. Air pollution included particulate matter pollution which consisted of ambient particulate matter pollution, household air pollution from solid fuels and ambient ozone pollution. Diabetes mellitus in this analysis included both type 1 and type 2 diabetes. Depression for the purpose of this analysis included depressive disorders – Major depressive disorder and Dysthymia. No definitive diagnosis criteria or harmonization methods are used in the original data sources and therefore, GBD-determined definitions of the listed risk factors and conditions were used in the present analysis.

### Data and statistical analysis

We examined point estimate numbers (with 95% uncertainty intervals, UI) for crude and age-standardized death rates, incidence rates and DALY rates for dementia in Afghanistan, Bangladesh, Bhutan, India, Maldives, Nepal, Pakistan and Sri Lanka. South Asia and global average estimates were also calculated for comparison purposes. Sex-specific and aggregated data were both examined. We also used GBD 2019 data to examine rates of risk factors related to chronic disease in South Asian countries – low physical activity, tobacco use, alcohol use, obesity, hypertension, diabetes mellitus, depression, hearing loss, and air pollution – that previous studies have identified as relevant risk factors for dementia [34]. All ages and age-standardized DALY rates data pertaining to all-cause contribution of these risk factors were extracted. The contribution and distribution of these risk factors for each South Asian nation was then computed. Linear regression analysis was employed to examine the relationships between the risk factors (independent variables) and dementia (dependent variable). Assumptions for linearity and normality of data were examined. Pearson correlation coefficients (R-values) and statistical significance (p-values) were computed. p<0.05 was considered statistically significant. All data were analyzed using python programming language.

## RESULTS AND DISCUSSION

### Population and aging statistics, and dementia as a health priority in South Asia

India was the most populated country in South Asia at the time of this analysis, with approximately 1.4 billion people, followed by Pakistan and Bangladesh **(Table 1**). Maldives was the least populated but most densely populated South Asian country. Dementia is strongly correlated with advanced age, therefore, life expectancies and proportion of older adults in different countries were examined. Life expectancy at birth was highest in Maldives (77.4 years) followed by Sri Lanka (76.8 years). Life expectancy at birth was lowest in Afghanistan (54.4 years), likely due to high maternal mortality rates, high prevalence of infectious disease and armed conflicts [35]. The percentage of older adults (individuals who were 65 years or older) was highest in Sri Lanka (12.4%), almost two-times higher than other South Asian countries. The lowest percentage of older adults was in Afghanistan (2.9%).

**Table 1.** South Asia statistics on populations and demographics. Data source: US Census Bureau international database (2024). Population density is listed in number of people per km^2^. Life expectancy at birth is listed in years.

| Country | Total population | Population density | Life expectancy at birth | Dependency ratio, 65+ years | % of 65+ years |
| --- | --- | --- | --- | --- | --- |
| Afghanistan | 40,121,552 | 61.5 | 54.4 | 5 | 2.9 |
| Bangladesh | 168,697,184 | 1296 | 75.2 | 11.6 | 7.8 |
| Bhutan | 884,546 | 23 | 73.7 | 9.5 | 6.7 |
| India | 1,409,128,296 | 473.9 | 68.2 | 9.9 | 6.8 |
| Maldives | 388,858 | 1304.9 | 77.4 | 8.6 | 6.1 |
| Nepal | 31,122,387 | 217.1 | 73 | 9.4 | 6.4 |
| Pakistan | 252,363,571 | 327.4 | 70.3 | 8.1 | 4.9 |
| Sri Lanka | 21,982,608 | 340.1 | 76.8 | 19.1 | 12.4 |

Setting and following a national-level agenda to assess dementia is an essential component of improving outcomes related to it [34]. Collectively, most LMICs in the world have poor or non-existing policies related to dementia. Compared to other regions of the world, there is limited investment into brain aging and dementia initiatives in South Asia. No countries in South Asia currently have a national plan on dementia. Only 4 South Asian countries (Bangladesh, India, Maldives, Pakistan) out of the 8 currently participate in the WHO Global Dementia Observatory **(Table 2)**. Even in the 4 countries that participate in the WHO Global Dementia Observatory, no dementia data had been compiled in a report between the period of 2017-2019. The status of other areas related to dementia awareness, dementia risk reduction, diagnosis, treatment and care, dementia carer support, dementia information systems and research is also similar across these countries highlighting an urgent need for action. Prior reports indicate that there are barriers to dementia assessment and care – stigma about dementia, poor patient engagement, limited trained healthcare professionals who can diagnose and manage dementia, competing healthcare system priorities and insufficient funding [36]. Studies have also identified the importance of establishing research priorities to improve outcomes related to dementia [37].

**Table 2.** Dementia as a health priority in South Asia. Data source: WHO Global Dementia Observatory (2019).

| Country | Included in/ contributes to WHO Global Dementia Observatory? | Availability and status of dementia reporting in the past 2 years, 2017-2019 |
| --- | --- | --- |
| Afghanistan | No | N/A |
| Bangladesh | Yes | No dementia data have been compiled in a report |
| Bhutan | No | N/A |
| India | Yes | No dementia data have been compiled in a report |
| Maldives | Yes | No dementia data have been compiled in a report |
| Nepal | No | N/A |
| Pakistan | Yes | No dementia data have been compiled in a report |
| Sri Lanka | No | N/A |

Research priorities identified are focused on prevention, reduction of dementia risk, delivery and quality of life and care for people with dementia and their caregivers. Other priorities identified were related to diagnosis, biomarkers, treatment development, basic research into disease mechanisms and public awareness of dementia. The South Asian region appears to lack in a majority of these areas owing to poor prioritization of dementia as a public health concern.

### Incidence, deaths and DALYs from all-cause dementia in South Asia

Using GBD 2019 data, crude and age-standardized rates of dementia in South Asian nations were considered **(Table 3**, **Figure 1)**. The crude death rates in South Asian countries were lower than the global average (20.98 per 100,000) **(Figure 1A)**. Amongst South Asian nations, Sri Lanka showed the highest death rate (18.95 per 100,000). This was much higher than the South Asia mean (9.48 per 100,000). Age-standardized death rates in some South Asian countries (Afghanistan and Sri Lanka) were higher than the South Asia and global average **(Figure 1B)**. Within South Asian countries, Afghanistan showed the highest age-standardized deaths (30.79 per 100,000) followed by Sri Lanka (23.53 per 100,000). Incidence rates of dementia in South Asia showed similar patterns. Crude incidence rate was highest in Sri Lanka (84.32 per 100,000) which was slightly lower than the estimated global average (93.52 per 100,000) **(Figure 1C)**. Age-standardized incidence rate was highest in Afghanistan (109.39 per 100,000) which was much higher than the global average (94.99 per 100,000), followed by Sri Lanka (86.29 per 100,000) **(Figure 1D**). Maldives also showed a relatively high age-adjusted incidence rate of dementia (86.00 per 100,000). For DALYs, highest crude rates were seen in Sri Lanka (321.52 per 100,000) which was slightly lower than the global average (326.68 per 100,000) **(Figure 1E)**. The highest age-standardized DALY rates were observed in Afghanistan (432.72 per 100,000) followed by Sri Lanka (342.98 per 100,000) **(Figure 1F)**. Both of these were higher than the global age-standardized DALY rates (338.68 per 1500,000). Prevalence rates were not examined in depth in the present study. GBD 2019 estimates show the highest crude prevalence for Sri Lanka 0.63% which is comparable to the global prevalence (0.69%).

**Figure 1.**
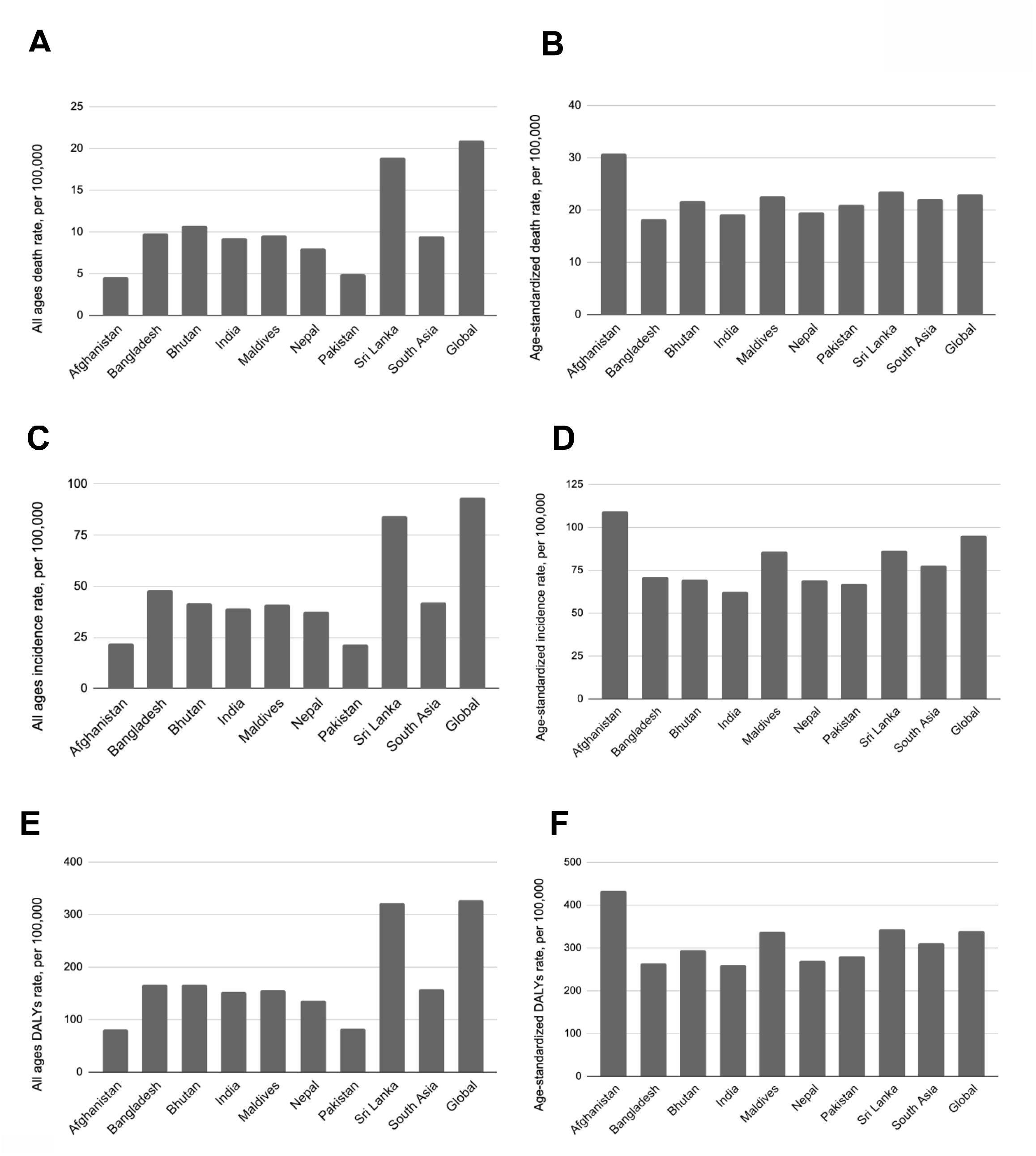
All ages (crude) **A)** death rates, **C)** incidence rates, **E)** DALY rates and age-standardized **B)** death rates, **D)** incidence rates, **F)** DALY rates of dementia in South Asian nations. South Asia and global averages are also shown for comparison. All rates are per 100,000. Disability-adjusted life years, DALYs. Data source: Global Burden of Disease Study (2019).

**Table 3.** All ages (crude) and age-standardized death, incidence and DALY rates for all-cause dementia in South Asian countries with uncertainty intervals (UI). Global averages are also shown for comparison. Data source: Global Burden of Disease Study (2019).

| Country | Death rate (per 100,000) |  | Incidence rate (per 100,000) |  | DALYs rate (per 100,000) |  |
| --- | --- | --- | --- | --- | --- | --- |
|  | *All ages* | *Age-standardized* | *All ages* | *Age-standardized* | *All ages* | *Age-standardized* |
| **Afghanistan** | 4.64 (1.10-12.54) | 30.79 (7.46-82.34) | 22.10 (18.56-25.57) | 109.39 (92.94-125.60) | 80.83 (33.26-192.81) | 432.72 (179.01-1001.95) |
| **Bangladesh** | 9.80 (2.38-27.25) | 18.23 (4.45-49.29) | 48.14 (41.10-55.88) | 71.28 (61.06-82.14) | 166.64 (72.32-377.64) | 263.10 (114.29-589.70) |
| **Bhutan** | 10.68 (2.56-28.70) | 21.68 (5.25-57.41) | 41.53 (35.44-47.99) | 69.41 (59.22-79.62) | 166.28 68.60-387.64) | 294.56 (120.22-672.40) |
| **India** | 9.28 (2.24-25.90) | 19.12 (4.57-52.01) | 38.93 (33.37-44.84) | 62.29 (53.08-71.43) | 152.84 (61.32-367.97) | 260.26 (103.52-616.93) |
| **Maldives** | 9.57 (2.36-24.66) | 22.70 (5.59-58.33) | 41.34 (35.50-47.11) | 86.00 (73.41-98.08) | 156.74 (66.96-351.04) | 338.10 (145.14-745.14) |
| **Nepal** | 8.05 (1.87-22.50) | 19.51 (4.53-53.31) | 37.78 (32.57-43.38) | 68.96 (58.69-79.24) | 136.76 (57.06-322.81) | 270.81 (111.54-636.11) |
| **Pakistan** | 4.89 (1.18-14.23) | 20.98 (5.01-58.43) | 21.75 (18.70-24.90) | 67.22 (57.19-77.28) | 82.02 (32.81-200.65) | 280.19 (110.44-683.03) |
| **Sri Lanka** | 18.95 (4.47-49.76) | 23.53 (5.52-60.92) | 84.32 (72.77-96.48) | 86.29 (73.91-98.61) | 321.52 (132.12-736.58) | 342.98 (140.10-771.73) |
| **South Asia** | 9.48 (2.27-25.70) | 22.07 (5.30-59.00) | 41.98 (36.00-48.30) | 77.61 (66.20-89.00) | 157.95 (65.56-367.14) | 310.34 (128.03-714.62) |
| **Global** | 20.98 (5.27-54.36) | 22.92 (5.83-59.20) | 93.52 (80.35-106.40) | 94.99 (81.59-107.86) | 326.68 (144.81-705.12) | 338.64 (151.02-731.27) |

It is important to highlight the absence of definitive diagnosis criteria leading to underreporting when compiling large amounts of global health data from various sources in the GBD project. For example, dementia is typically diagnosed using cognitive testing (e.g., Montreal Cognitive Assessment, Mini Mental State Examination), brain imaging (e.g., magnetic resonance imaging) blood or cerebrospinal fluid (CSF) tests (e.g., amyloid β or p-tau levels) and others. It is difficult to ascertain which method was used in aggregated data. Due to absence of technology and infrastructure, brain MRI or blood/ CSF tests might not be available in many low-income settings so it is likely that cognitive and neurophysiological evaluations are prioritized but they may have limited sensitivity in case detections. Therefore, these broad estimates must be followed up with in-depth studies in order to accurately quantify dementia levels in each population. Previous independent reports that assessed dementia rates in locally-conducted community-based cohort studies show prevalence rates in South Asian nations – 8% in Bangladesh [38], 7.4% in India [39], and 4% in Sri Lanka according to a report from 2003 [40].

Sex-specific analysis showed that the burden of dementia in South Asian countries was higher amongst females than males (data not shown). This is an observation that has been made elsewhere in the world as well where females are disproportionately affected by the disease [41]. Mechanisms behind this are not fully understood but differential life course exposures and metabolic changes that occur during midlife, and menopause appear to play a role [42] [43]. Females also have longer lifespans on average compared to males which may further contribute to this.

Overall, within South Asian countries, the highest crude rates of deaths, incidence and DALYs related to dementia were seen in Sri Lanka while the highest age-standardized rates were observed in Afghanistan, followed by Sri Lanka and Maldives. These findings indicate that population aging likely plays a substantial role in the dementia burden in South Asia similar to other parts of the world [44]. Previous studies have shown that aging is the strongest predictor of dementia [45], and findings from this analysis also emphasize this within the context of South Asia. As a result, countries with higher proportions of older adults in this age range may experience a disproportionately higher burden of dementia. Age-specific analyses showed an age-dependent increase in dementia deaths, incidence and burden rates in all 8 South Asian countries as well as globally (data not shown). Dementia rates were higher in younger age groups in Sri Lanka, compared to Afghanistan (data not shown). Overall, this may also contribute to higher crude dementia rates observed in Sri Lanka. However, the total rates of dementia were much higher in older age groups across all countries.

There also may be other factors that can contribute to the high dementia burden, in addition to population aging. A recent study showed that multidimensional poverty is associated with dementia in Afghanistan [46]. The main contributors identified during this study were education, employment, health and living conditions. Prior research has shown that trauma is linked to poor mental health outcomes, e.g., post-traumatic stress disorder and dementia amongst US Veterans [47]. Whether this phenomena specifically applies to countries in South Asia that have been in armed conflicts - Afghanistan and Sri Lanka, is currently unknown. Another consideration is how countries are geographically located, and are categorized as a result.

Afghanistan is geographically close to the Middle East and North African (MENA) region, and is often considered a middle eastern nation according to some classifications, including the GBD project. Studies have shown that the prevalence of dementia is very high in the MENA region [48]. How these geographical factors contribute to dementia data of individual countries, particularly where data are sparse or missing, can only be fully ascertained by examining high-quality, country-specific data in rigorous follow-up studies [49].

### Etiological factors driving dementia in South Asia

We interrogated GBD 2019 study data to ascertain whether there were any dementia risk factors that were prevalent in South Asian countries. There are a few known risk factors of dementia – air pollution, alcohol use, high BMI, high fasting blood glucose, high systolic blood pressure, low physical activity, tobacco, hearing loss, depressive disorders and diabetes mellitus, amongst others. The GBD 2019 study identified smoking, high body-mass index and high fasting plasma glucose as the major risk factors contributing to global dementia burden [21]. We examined crude and age-standardized DALY rates for each risk factor available through the GBD 2019 study for each South Asian nation. There were appreciable similarities as well as differences in crude and age-standardized DALYs associated with various risk factors within different South Asian nations. High systolic blood pressure and high fasting blood glucose appeared to be notable contributors for the overall burden across South Asian populations **(Figure 2)**. In some nations, other risk factors were more prominent. For example, air pollution made up a large proportion of the contributing risk in Nepal, India and Bangladesh while Diabetes Mellitus was disproportionately higher in Sri Lanka. These findings indicate that while there might be common drivers relevant to dementia in different South Asian nations, the burden might be modified by country-specific, unique risk factors as well. The exact contribution of each risk factor to dementia burden specifically was not analyzed here as the goal was to compare the prominence of well-established dementia risk factors in each South Asian nation.

**Figure 2.**
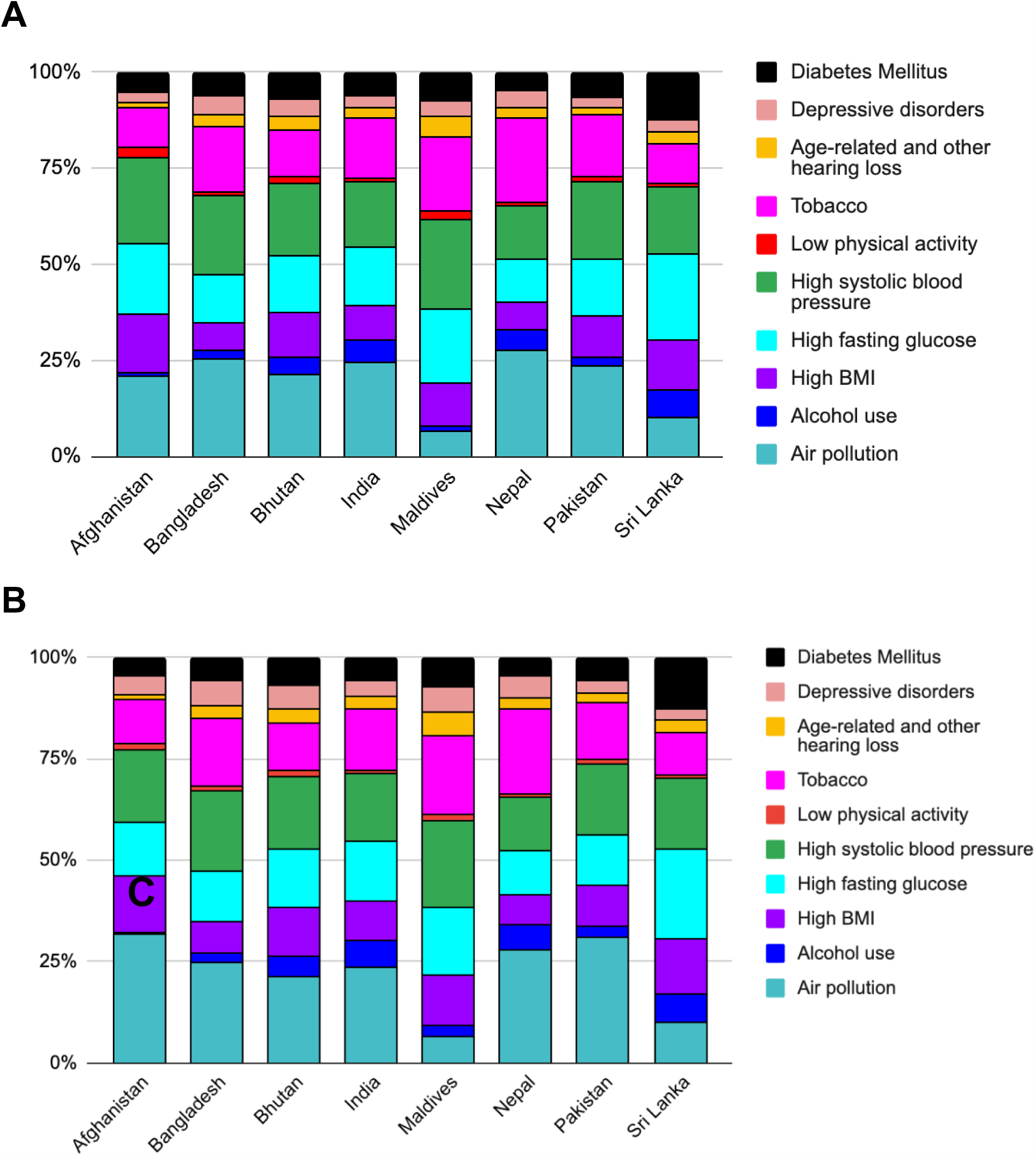
A) All ages (crude) and **B)** age-standardized DALY rates of known disease risk factors that have implications for dementia, compared between the 8 South Asian nations. Data source: BMI, body mass index. Global Burden of Disease Study (2019).

In order to understand the relationship between these risk factors and dementia within South Asia, a linear regression analysis was conducted where Pearson correlation coefficients (R-value) and p-values were computed. For crude DALYs, the strongest positive significant correlations for dementia were observed with age-related and other hearing loss (R=0.87, p=0.01), Diabetes Mellitus (R=0.85, p=0.01) and high fasting blood glucose (R=0.74, p=0.04) (**Table 4**). A moderate correlation was observed for alcohol use (R=0.67, p=0.07) which was not statistically significant. For age-standardized DALYs, the strongest correlations for dementia were seen with low physical activity (R=0.78, p=0.02) and high BMI (R=0.78, p=0.02) and high fasting blood glucose (R=0.77, p=0.03). In addition, high systolic blood pressure (R=0.64, p=0.09) and Diabetes Mellitus (R=0.52, p=0.19) also showed noteworthy correlations which were not statistically significant. An important point to note is that many of these risk factors also exhibit strong correlations with each other (data not shown), which can have a direct bearing on the results. For example, high fasting glucose and Diabetes Mellitus are very strongly correlated (R=0.97, crude rate and R=0.78, age-standardized rate). Therefore, ascertaining the exact causal role of a given risk factor on dementia is not possible through an analysis of this nature. Future analyses should consider this, where advanced statistical models may be more useful.

**Table 4.** Correlation between biological, behavioral and environmental risk factors and dementia in South Asia. Pearson correlation coefficients (R-values) and statistical significance (p-values) are shown for crude and age-standardized DALYs. *Statistically significant (p<0.05). Data source: Global Burden of Disease Study (2019).

| Risk factor | Correlation coefficient (Crude DALYs) | p-value (Crude DALYs) | Correlation coefficient (age-standardized DALYs) | p-value (age-standardized DALYs) |
| --- | --- | --- | --- | --- |
| Air pollution | -0.63 | p=0.093 | 0.06 | p=0.885 |
| Alcohol use | 0.67 | p=0.068 | -0.49 | p=0.222 |
| High BMI | 0.39 | p=0.340 | 0.78* | p=0.022 |
| High fasting blood glucose | 0.74* | p=0.037 | 0.77* | p=0.026 |
| High systolic blood pressure | 0.28 | p=0.498 | 0.64 | p=0.090 |
| Low physical activity | -0.28 | p=0.501 | 0.78* | p=0.021 |
| Tobacco | -0.15 | p=0.719 | -0.18 | p=0.664 |
| Age-related and other hearing loss | 0.87* | p=0.005 | -0.50 | p=0.205 |
| Depressive disorders | -0.35 | p=0.398 | 0.01 | p=0.989 |
| Diabetes Mellitus | 0.85* | p=0.007 | 0.52 | p=0.187 |

Collectively, the findings from this study align closely with other studies showing that metabolic risk factors, particularly hyperglycemia, play an important role in dementia incidence and progression [50][51]. Preventing and controlling metabolic dysfunction should therefore be an urgent priority for South Asians in order to prevent and control a host of chronic diseases, including dementia [52].

Other studies have also shown that of some of these metabolic risk factors are abundantly prevalent in some South Asian countries. A recent publication by an independent group showed that the prevalence of type 2 diabetes amongst Sri Lankans is 23% [53]. The same group showed that the prevalence of prediabetes is 30.5%. These levels are significantly higher than the global average and other countries in the Asia Pacific region [54]. The exact reasons for the high prevalence of metabolic risk factors in South Asian countries is not fully understood but poor diets, lack of physical activity, genetic predispositions and life course factors, e.g., biological programming may all be contributing [55].

However, it is important to note that there are several other dementia risk factors that were not considered here. For example, low educational attainment has been associated with poorer dementia outcomes in previous studies [56]. Vision loss and muscle strength are also emerging as predictors of dementia [57][58]. Even though some of these data are not challenging to collect, many of them are not been considered in population-level studies leading to missed opportunities to better understand dementia determinants. There are other risk factors that are more costly to measure and therefore might not be feasible to measure in low income and population-level settings. For example, genetic factors such as apolipoprotein E ε4 (APOE ε4) is also strongly linked with dementia incidence and progression [59] but requires laboratory-based genotyping. Due to absence of appropriate current data, these factors could not be considered in the present analysis.

### The need for more data

The GBD Study is a useful resource to identify global health trends and patterns that can help drive policy focus [49]. These data are largely based on estimates arrived at through various statistical models. These data are also compiled from thousands of different sources which may lead to underreporting, and missing data for some countries. Furthermore, some conditions are aggregated together in the GBD which may affect the findings presented here. For example, in the GBD, hearing loss that is aging-related and other causes-related are combined. Aging-associated hearing loss is a strong predictor of dementia but by merging these data with other types of hearing loss, some trends may be lost. In the absence of data collected through local studies, which is often the case with LMIC countries, sources such as the GBD provide valuable starting points for health trend analyses. However, the need for further research on unique factors that may drive dementia and also trajectories of dementia trends in individual countries is pressing. For example, there is also very limited data on mild cognitive impairment (MCI) and how cognitive decline progresses as individuals age in South Asia. Furthermore, there’s also poor understanding of the different types of dementia in the region. The paucity of reliable data from this region has been previously noted for many years [60].

### Considerations for future studies

Findings from broad, large global health projects must be followed up using more specific analyses that focus on interrogating observations made. Several unique challenges must be overcome when planning and executing longitudinal follow-up studies aiming to characterize brain aging and dementia in South Asian countries. These include financial, cultural and structural barriers. **Figure 3** highlights major considerations applicable to countries in South Asia, as well as others that are LMIC. Most countries in South Asia have limited medical research funding, which poses a major problem for establishing state-funded programs that are sustainable and continuous. However, overcoming these initial barriers through strategic partnerships, can help set up powerful data and biological sample repositories that can also serve as strong platforms for research, training, education and public awareness. Establishing standard definitions and diagnostic criteria for dementia as well as utilizing biomarkers, both behavioral and imaging are important. These long-term studies can also add immense value to overall understanding of dementia and risk factors, ultimately paving the way for prevention and therapeutic strategies that encompass global populations.

**Figure 3.**
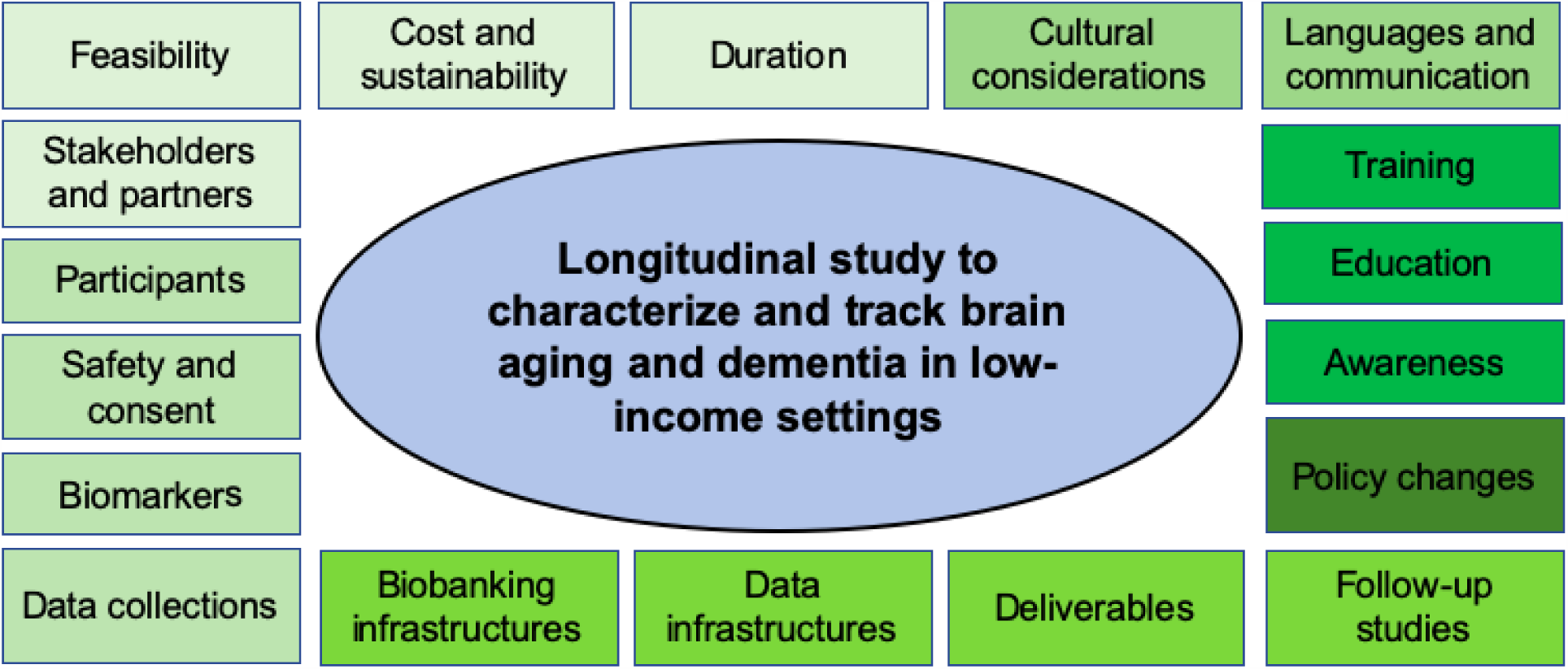
Considerations for setting up future longitudinal studies examining brain aging and dementia in low-income settings such as countries in South Asia.

Such follow-up, focused studies have shown that dementia rates in some South Asian countries have historically been underestimated due to underdiagnoses and underreporting. For example, a recent report showed that there are a few million more cases of dementia in India than previously estimated [39]. This is likely the case for many other LMICs that don’t have dedicated resources to track and monitor the status of aging-associated conditions like dementia. This can be exacerbated by classifying dementia as a normal part of the aging process. It is also noteworthy that the COVID-19 pandemic has negatively affected diagnosis, management and research related to dementia in many South Asian countries. Additionally, country-specific concerns have also significantly worsened the dementia burden. For example, Sri Lanka was hit by an economic crisis in 2022, leading to serious financial and societal issues that have invariably affected the health sector. The exact magnitude of this crisis and its effects on particularly human health outcomes is unknown.

There are also specific cultural, religious and language factors that must be considered when examining dementia burden in South Asia. Most South Asian countries promote strong family structures, and therefore, have multi-generational households where older adults live with younger individuals. When older adults are diagnosed with dementia, caretaking responsibilities may be taken up by younger members of the family due to value systems. Not being adequately equipped and trained to care for a dementia patient can lead to a significant caregiver burden [61][62]. South Asian countries also do not have formal caregiver structures in place, e.g., nursing homes, social workers, community-based healthcare teams, which are essential to effectively reducing overall burden of dementia. Reliable data on caregiver systems are also sparse in South Asian countries and are not addressed in large studies such as the GDB. The present study did not examine dementia burden amongst South Asians who live in other parts of the world [63]. In spite of dementia outcomes being better in high income countries, South Asians living in these parts of the world may experience bias and discrimination when seeking dementia-related care [64]. There’s also dementia-related stigma within South Asian communities which must be addressed [65].

With rapidly growing older adult populations, these concerns are expected to magnify and countries that are not prepared will likely experience the heaviest financial, societal and public health burden. Efforts dedicated to accurately measuring aging and brain aging biomarkers, and also dementia statistics and determinants are vital first steps that must be undertaken in order to understand the true situation of dementia in these countries. Through this, contributing risk factors can be identified so that they can be controlled, where possible, ultimately leading to improved outcomes related to dementia. Follow-up studies can also help facilitate the a better understanding of dementia subtypes, timelines, and molecular targets that may be critical for identifying therapeutic interventions in the future.

## CONCLUSIONS

In this preliminary summary analysis using publicly available dementia data, we highlight the status and determinants of dementia burden in 8 South Asian nations. Our findings highlight that South Asian countries are lagging behind other regions with regard to prioritization of brain aging and dementia as healthcare and research items that require urgent attention. Sri Lanka (highest proportion of older adults in South Asia) and Afghanistan (lowest proportion of older adults in South Asia) are experiencing the most significant burden of dementia, driven by population aging and other factors, respectively. We also identify that metabolic dysfunction strongly contributes to the dementia burden in South Asian countries. Overall, this analysis emphasizes the need for country-specific, longitudinally-collected data on risk factors and biomarkers of dementia in South Asia. Therefore, prioritizing research, awareness and services related to dementia can help South Asian countries prepare for the impending public health challenges related to population aging.

## DATA AVAILABILITY AND SHARING

All raw data can be downloaded from primary, publicly available data sources listed in the manuscript. All analyzed data and codes used for analysis are available upon request from corresponding author.

## AUTHOR CONTRIBUTIONS

DLP; Planning the study - DLP, NP; Compiling, curating and analyzing data - DLP, NP, SL, CVLT, NMA, US, CA, SJ, CD; Discussion, data interpretation, proofreading and editing - DLP; Writing original draft and supervision.

## ACKNOWLEDGEMENTS

The authors would like to acknowledge the primary data sources – US Census, WHO Global Dementia Observatory and the Institute for Health Metrics and Evaluation (IHME). We are also grateful to Professor Vajira HW Dissanayake, Dean of University of Colombo Faculty of Medicine for his support and Texas Tech University Innovation Hub for their ongoing assistance of our work.

## FUNDING STATEMENT

No funding sources to declare

## COMPETING INTERESTS

None

## Notes

### Competing Interest Statement

The authors have declared no competing interest.

### Author Declarations

All data used in the study were obtained from public sources and are listed in the manuscript.

